# Evaluation of the diagnostic value of YiDiXie™-SS, YiDiXie™-HS and YiDiXie™-D in gastric cancer

**DOI:** 10.1101/2024.08.21.24312336

**Authors:** Huimei Zhou, Chen Sun, Yanrong Qi, Yutong Wu, Xutai Li, Zhenjian Ge, Wenkang Chen, Yingqi Li, Pengwu Zhang, Shengjie Lin, Wuping Wang, Siwei Chen, Wei Li, Xi Li, Ling Ji, Yongqing Lai

**Affiliations:** Department of Urology, Peking University Shenzhen Hospital, Shenzhen, 518036; Institute of Urology, Shenzhen Peking University-The Hong Kong University of Science and Technology Medical Center, Shenzhen, 518036; The Fifth Clinical Medical College of Anhui Medical University, Hefei 230032; Department of Gastroenterology, Haibin People’s Hospital of Tian Jin Binhai New Area, Tianjin, 300280; Shantou University Medical College, Shantou, Guangdong 515041; Shenzhen University Health Science Center, Shenzhen, China 518055; Shenzhen KeRuiDa Health Technology Co., Ltd., Shenzhen, 518071; Department of Gastroenterology, Peking University Shenzhen Hospital, Shenzhen, 518036; Department of Laboratory Medicine, Peking University Shenzhen Hospital, Shenzhen, 518036

**Keywords:** Gastric cancer, Fecal occult blood test, CEA, CA125, CA19-9, False-positive, False-negative, YiDiXie ™ -SS

## Abstract

**Background:** Gastric cancer poses a severe risk to public health and has a substantial financial impact. Tumor markers such as CEA, CA125, CA19-9, and others, as well as the fecal occult blood test (FOBT), are frequently utilized for gastric cancer screening and initial diagnosis. However, false-positive results of FOBT, CEA, CA125, and CA19-9 can lead to misdiagnosis and erroneous gastroscopy, while their false-negative results can lead to missed diagnosis and delayed treatment. Finding practical, affordable, and non-invasive diagnostic techniques is crucial to lowering the false-positive and false-negative rates of FOBT and other markers. The aim of this study was to evaluate the diagnostic value of YiDiXie ™-SS, YiDiXie™-HS and YiDiXie™-D in gastric cancer.

**Patients and methods:** This study included 602 subjects (Malignant group, n=222; Benign group, n=380 cases). The remaining serum samples of the subjects were collected and the sensitivity and specificity of the YiDiXie™-SS, YiDiXie™ -HS and YiDiXie™-D were evaluated using the YiDiXie™ all-cancer detection kit.

**Results:** The sensitivity of YiDiXie™-SS was 99.5% (95% CI: 97.5% - 100%) and its specificity was 64.5% (95% CI: 59.5% - 69.1%). This means that YiDiXie™-SS has an extremely high sensitivity and relatively high specificity in gastric tumors.YiDiXie™-HS has a sensitivity of 96.8% (95% CI: 93.6% - 98.5%) and a specificity of 89.5% (95% CI: 86.0% - 92.2%). This means that YiDiXie™ -HS has high sensitivity and high specificity in gastric tumors. The sensitivity of YiDiXie™-D was 83.3% (95% CI: 77.9% - 87.7%) and its specificity was 95.5% (95% CI: 93.0% - 97.2%). This means that YiDiXie™-D has relatively high sensitivity and very high specificity in gastric tumors. YiDiXie™-SS significantly reduced the false-positive rates of FOBT, CEA, CA125, and CA19-9 with essentially no increase in malignancy leakage.YiDiXie™-HS substantially reduced the false-negative rates of FOBT, CEA, CA125, and CA19-9. YiDiXie™-D significantly reduces the false positive rate of FOBT, CEA, CA125, CA19-9. YiDiXie™-D significantly reduces the false negative rate of FOBT, CEA, CA125, CA19-9 while maintaining a high level of specificity.

**Conclusion:** YiDiXie™-SS has very high sensitivity and relatively high specificity in gastric tumors.YiDiXie™-HS has high sensitivity and high specificity in gastric tumors.YiDiXie ™ -D has relatively high sensitivity and very high specificity in gastric tumors. YiDiXie ™ -SS significantly reduces the false-positive rates of FOBT, CEA, CA125, and CA19-9 with essentially no increase in delayed treatment of gastric cancer.YiDiXie™-HS significantly reduces the false-negative rates of FOBT, CEA, CA125, and CA19-9.YiDiXie™-D can significantly reduce the false-positive rate of FOBT, CEA, CA125 and CA19-9, or significantly reduce the false-negative rate of FOBT, CEA, CA125 and CA19-9 while maintaining a high degree of specificity. YiDiXie™ test has an important diagnostic value in gastric cancer, and is expected to solve the problems of “high false positive rate” and “high false negative rate” of FOBT, CEA, CA125 and CA19-9.

## INTRODUCTION

Gastric cancer is one of the most common malignant tumors. In 2022, there were more than 968,000 new cases of gastric cancer and nearly 660,000 deaths, ranking among the highest in the world in terms of morbidity and mortality^1^. Over the past half century, gastric cancer incidence and mortality have been high in most populations^2-3^. Recent studies have shown an increasing incidence in younger age groups, especially in low-incidence populations^4-5^. The overall survival rate of early gastric cancer (GC) can exceed 96% whether it is treated by endoscopy or surgery^6^, but most cancer cases are diagnosed at an advanced stage. The median survival rate in the late stage is less than 12 months^7^. The 5-year survival rate in other European and American countries fluctuates in the range of 15%-30%, except for Japan, which is relatively high^8-9^. Gastric cancer, as a highly aggressive malignancy, remains a global health problem today^10^.

On the one hand, due to the convenience, Fecal occult blood test (FOBT) and CEA, CA125, CA19-9 and other tumor markers are widely used in the screening or preliminary diagnosis of gastric cancer. For FOBT positive patients, the prevalence of FOBT positive upper digestive tract lesions was as common as that of lower digestive tract lesions^11^. The source of FOBT positive rates in the upper digestive tract alone is mainly determined by the prevalence of gastric cancer and Helicobacter pylori^12-14^. H. pylori may cause a higher incidence of gastrointestinal ulcers or mucosal inflammation, and 1/3 of FOBT-positive patients have normal EGD, most patients who received EGD were found to have inflammation of the gastrointestinal mucosa: non-erosive gastritis (30.7%), duodenitis (9.8%), esophagitis (4.9%) and erosive gastritis (1.7%)^15^. Therefore, there are many reasons for positive FOBT results, and gastric cancer is only one of them, resulting in a high false-positive rate of FOBT for gastric cancer. As traditional tumor markers, CEA, CA125 and CA19-9 are expressed in a variety of cancers and even benign diseases. Therefore, the specificity of gastric cancer diagnosis is not high, It also produces a high false positive rate^16-20^. Studies have shown that the false-positive rate of CEA for gastric cancer diagnosis is as high as 33%^21^. When FOBT, CEA, CA125, CA19-9 and other markers are positive, patients usually undergo endoscopy and biopsy^22-24^. False-positive results of FOBT and other markers mean that patients have received unnecessary endoscopy, and patients may face adverse consequences such as infection, mental distress, expensive examination costs, and examination injuries. Therefore, there is an urgent need to find a convenient, economical and non-invasive diagnostic method to reduce the false-positive rate of FOBT and other markers.

On the other hand, because early gastric cancer usually has no symptoms, tests such as FOBT do not detect early stomach cancer, and the sensitivity is extremely low. By the time stomach cancer is diagnosed because it is positive for FOBT, most stomach cancers are advanced and difficult to cure^25^. The positive rates of the same four markers of early gastric cancer (CEA, CA125, CA19-9) varied greatly. The positive rate of CEA has been reported to be 4.3% to 15.4%^26-29^. The positive rate of CA19-9 was 4.8%-11.7%^28-29^, and the positive rate of CA125 was 1.9%-6.7%^29-30^. Even the combined positive rate of multiple markers was only 10.4%^29^. However, only very few tumor markers that should be diagnosed as positive are correctly diagnosed, with a false-negative rate of more than 90%, which means that even the combination of tumor markers in the diagnosis of early gastric cancer is extremely low. When FOBT and other markers are negative, patients are usually observed and regularly followed up, and only in areas with high incidence of gastric cancer will endoscopic screening be performed again^22-24^. False-negative results of FOBT and other markers mean that malignant tumors are misdiagnosed as benign diseases, which may lead to delayed treatment, the progression of malignant tumors, and even the development of advanced stages. Therefore, patients will have to bear the adverse consequences of poor prognosis, high treatment costs, poor quality of life, and short survival. Therefore, it is urgent to find a convenient, economical and non-invasive diagnostic method to reduce the false-negative rate of FOBT and other markers.

Based on the detection of novel tumor markers of miRNA in serum, Shenzhen KeRuiDa Health Technology Co., Ltd. has developed an in vitro diagnostic test product YiDiXie ™ all-cancer test (the YiDiXie™ test).^31^ With just 200μl of whole blood or 100 μl of serum at a time, many cancer types can be detected^31^. the YiDiXie™ test includes three products with different performance: YiDiXie ™-HS, YiDiXie™-SS, and YiDiXie™-D^31^.

The purpose of this study was to evaluate the diagnostic value of YiDiXie™-SS, YiDiXie™-HS and YiDiXie™-D in gastric cancer.

## PATIENTS AND METHODS

### Study design

This work is part of the SZ-PILOT study (ChiCTR2200066840), “Evaluating the value of the YiDiXie™ test as an adjunct diagnostic in multiple tumors”.

The SZ-PILOT study (ChiCTR2200066840) was a single-center, prospective, observational study. Subjects who signed the pan-informed consent to donate the remaining samples at admission or physical examination were included, and 0.5ml of their remaining serum samples were collected for this study.

Blind method was used in this study. Neither the experimentor conducting the YiDiXie™ test nor the KeRuiDa laboratory technicians who determined the YiDiXie™ test results were aware of the study subjects’ clinical profiles. The results of the YiDiXie ™ test were also not known to the clinical experts who evaluated the subjects’ clinical data.

This study was approved by the Ethics Committee of Peking University Shenzhen Hospital and carried out in accordance with the International Coordination Conference on Quality Management of Drug Clinical Trials and the Declaration of Helsinki.

### Participants

Subjects with gastric cancer and gastric benign diseases with FOBT, CEA, CA125 and CA19-9 detection data were included in this study. Subjects in the two groups were separately enrolled, and all subjects meeting the inclusion criteria were continuously included.

This study initially included hospitalized patients with “suspected (solid or blood) malignancy” who signed a pan-informed consent to donate the remaining samples. Subjects with postoperative pathological diagnosis of “malignant tumor” were included in the malignant group, and those with postoperative pathological diagnosis of “benign disease” were included in the benign group. Ambiguous pathological results were excluded from this study. The benign group also included healthy checkers with colonoscopy results. Part of the samples from the malignant tumor group and healthy physical examination samples from the benign group were used in other previous studies of our group^31^.

Subjects whose serum samples were disqualified prior to the YiDiXie ™ test were excluded from the study. For specific information on inclusion and exclusion, refer to the previous articles of our group^31^.

### Sample collection, processing

The serum samples used in this study were taken from serum remaining after normal treatment and no additional blood was drawn. Approximately 0.5ml serum samples were collected from the remaining serum of medical laboratory subjects and stored at -80 °C for subsequent the YiDiXie™ test.

### The YiDiXie test

The YiDiXie ™ test was performed using YiDiXie ™ all-cancer detection kit. YiDiXie ™ all-cancer detection kit is an in vitro diagnostic kit developed and produced by Shenzhen KeRuiDa Health Technology Co., Ltd. for fluorescent quantitative PCR instrument^31^. It detects the expression levels of dozens of miRNA biomarkers in blood serum to determine whether cancer is present in the subject’s body.^31^ It pre-defines appropriate thresholds for each miRNA biomarker, thus ensuring that each miRNA biomarker has high specificity, and integrates these independent assays through a parallel assay model to significantly increase sensitivity and maintain higher specificity for broad-spectrum cancers.^31^

The YiDiXie™ test includes three different test products: YiDiXie™-HS, YiDiXie™-SS, and YiDiXie™-D. YiDiXie ™ -HS was developed with sensitivity and specificity in mind. YiDiXie ™ -SS significantly increases the number of miRNA trials to achieve extremely high sensitivity for all clinical stages of all malignancies. YiDiXie ™ -Diagnosis (YiDiXie ™ -D) dramatically increases the diagnostic threshold for a single miRNA test to achieve extremely high specificity for all malignancies.^31^

The YiDiXie™ test was performed according to the instructions for the YiDiXie ™ all cancer detection kit. For details, refer to the previous articles of our group^31^.

The original test results were analyzed by laboratory technicians of Shenzhen KeRuiDa Health Technology Co., Ltd., and the YiDiXie™ test results were determined to be “positive” or “negative”^31^.

### Extraction of clinical data

The clinical, pathological, laboratory and imaging data in this study were extracted from patients’ hospital records or physical examination reports. Clinical staging was evaluated by trained clinicians according to the AJCC Staging Manual (7th or 8th edition)^32-33^.

### Statistical analyses

For demographic and baseline characteristics, report descriptive statistics. For categorical variables, calculate the number and percentage of subjects in each category; For continuous variables, total number of subjects (n), mean, standard deviation (SD), or standard error (SE), median, first quartile (Q1), third quartile (Q3), minimum, and maximum are calculated. 95% confidence intervals (CI) were calculated for multiple biomarkers using the Wilson (score) method.

## RESULTS

### Participant disposition

This study involved a total of 602 participants (222 malignant cases and 380 benign cases). The demographic and clinical characteristics of the 602 study participants are shown in Table 1.

**Table 1.** Participants’ demographic and clinical manifestration.

|  | Malignant (n =222) | Benign (n =380) | Total (N = 602) |
| --- | --- | --- | --- |
| Age, years |  |  |  |
| Mean (SD) | 58.8 (12.79) | 52.0 (11.02) | 54.5 (12.14) |
| Median (Q1,Q3) | 60 (50, 67) | 53 (45, 60) | 55 (47, 63) |
| Min, max | 20, 87 | 23, 78 | 20, 87 |
| Age, group, n (%) |  |  |  |
| < 50 | 48 (21.6) | 128 (33.7) | 176 (29.2) |
| ≥ 50 | 174 (78.4) | 252 (66.3) | 426 (70.8) |
| < 65 | 140 (63.1) | 332 (87.4) | 472 (78.4) |
| ≥ 65 | 82 (36.9) | 48 (12.6) | 130 (21.6) |
| Sex, n (%) |  |  |  |
| Female | 70 (31.5) | 171 (45.0) | 241 (40.0) |
| Male | 152 (68.5) | 209 (55.0) | 361 (60.0) |
| Body mass index (kg/m2) |  |  |  |
| n | 219 | 341 | 560 |
| Mean (SD) | 22.3 (3.09) | 23.6 (3.34) | 23.1 (3.31) |
| Median (Q1,Q3) | 22.0 (20.2, 24.4) | 23.2 (21.3, 25.7) | 22.9 (20.8, 25.3) |
| Min, max | 14.3 29.8 | 15.4 35.2 | 14.3 35.2 |
| Body mass index category, n (%) |  |  |  |
| Underweight | 28 (12.6) | 14 (3.7) | 42 (7.0) |
| Benign | 130 (58.6) | 185 (48.7) | 315 (52.3) |
| Overweight | 52 (23.4) | 107 (28.2) | 159 (26.4) |
| Obese | 9 (4.1) | 35 (9.2) | 44 (7.3) |
| Missing | 3 (1.4) | 39 (10.3) | 42 (7.0) |
| AJCC clinical stage |  |  |  |
| Stage I | 70 (31.5) |  | 70 (31.5) |
| Stage II | 25 (11.3) |  | 25 (11.3) |
| Stage III | 51 (23.0) |  | 51 (23.0) |
| Stage IV | 72 (32.4) |  | 72 (32.4) |
| Missing | 4 (1.8) |  | 4 (1.8) |

Q1,Q3, first quartile, third quartile; SD, standard deviation.

Demographic and clinical variables were similar between the two research groups (Table 1). The average (standard deviation) age was 54.5 (12.14) years, with 40.0% (241/602) being female.

### Diagnostic performance of YiDiXie™-SS

As shown in Table 2, the sensitivity of YiDiXie™-SS was 99.5% (95% CI: 97.5% - 100%) and its specificity was 64.5% (95% CI: 59.5% - 69.1%). This means that YiDiXie™-SS has very high sensitivity and relatively high specificity in gastric tumors.

**Table 2.**
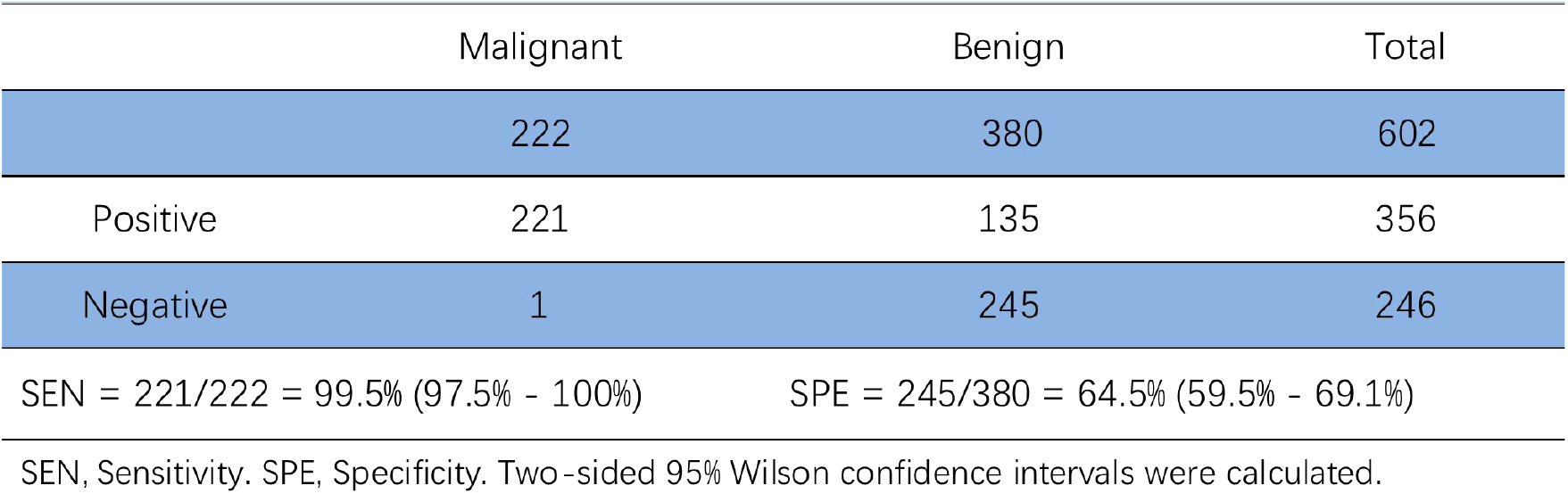
Performance of YiDiXie^™^-SS.

|  | Malignant | Benign | Total |
| --- | --- | --- | --- |
|  | 222 | 380 | 602 |
| Positive | 221 | 135 | 356 |
| Negative | 1 | 245 | 246 |

SEN, Sensitivity. SPE, Specificity. Two-sided 95% Wilson confidence intervals were calculated.

### Diagnostic performance of YiDiXie™-HS

As shown in Table 3, the sensitivity of YiDiXie™ -HS was 96.8% (95% CI: 93.6% - 98.5%) and its specificity was 89.5% (95% CI: 86.0% - 92.2%). This means that YiDiXie ™ -HS has high sensitivity and high specificity in gastric tumors.

**Table 3.** Performance of YiDiXie^™^-HS.

|  | Malignant | Benign | Total |
| --- | --- | --- | --- |
|  | 222 | 380 | 602 |
| Positive | 215 | 40 | 255 |
| Negative | 7 | 340 | 347 |

SEN, Sensitivity. SPE, Specificity. Two-sided 95% Wilson confidence intervals were calculated.

### Diagnostic performance of YiDiXie™-D

As shown in Table 4, the sensitivity of YiDiXie™-D was 83.3% (95% CI: 77.9% - 87.7%) and its specificity was 95.5% (95% CI: 93.0% - 97.2%). This means that YiDiXie™-D has relatively high sensitivity and very high specificity in gastric tumors.

**Table 4.** Performance of YiDiXie^™^-D.

|  | Malignant | Benign | Total |
| --- | --- | --- | --- |
|  | 222 | 380 | 602 |
| Positive | 185 | 17 | 202 |
| Negative | 37 | 363 | 400 |

SEN, Sensitivity. SPE, Specificity. Two-sided 95% Wilson confidence intervals were calculated.

### Diagnostic performance of YiDiXie™-SS in FOBT, CEA, CA125, CA19-9 positive patients

In order to solve the difficult problem of high false-positive rate of FOBT, CEA, CA125, CA19-9, YiDiXie™-SS was applied to FOBT, CEA, CA125, CA19-9 positive patients.

As shown in Table 5, YiDiXie™-SS significantly reduced the false-positive rates of FOBT, CEA, CA125, and CA19-9 with essentially no increase in malignant tumor leakage.

**Table 5.** Performance of different Items.

| Items | Total N | Positive | Sensitivity % (95% CI)<sup>a</sup> | Total N | Negative | Specificity % (95% CI)<sup>b</sup> |
| --- | --- | --- | --- | --- | --- | --- |
| YiDiXie™-SS in FOBT-positive | 13 | 13 | 100% (77.2% - 100%) | 4 | 3 | 75.0% (30.1% - 98.7%) |
| YiDiXie™-HS in FOBT-negative | 86 | 83 | 96.5% (90.2% - 99.0%) | 22 | 19 | 86.4% (66.7% - 95.3%) |
| YiDiXie™-D in FOBT-positive | 13 | 12 | 92.3% (66.7% - 99.6%) | 4 | 4 | 100% (51.0% - 100%) |
| YiDiXie™-D in FOBT-negative | 86 | 70 | 81.4% (71.9% - 88.2%) | 22 | 21 | 95.5% (78.2% - 99.8%) |
| YiDiXie™-SS in CEA-positive | 30 | 30 | 100% (88.6% - 100%) | 15 | 11 | 73.3% (48.0% - 89.1%) |
| YiDiXie™-HS in CEA-negative | 187 | 181 | 96.8% (93.2% - 98.5%) | 364 | 325 | 89.3% (85.7% - 92.1%) |
| YiDiXie™-D in CEA-positive | 30 | 29 | 96.7% (83.3% - 99.8%) | 15 | 14 | 93.3% (70.2% - 99.7%) |
| YiDiXie™-D in CEA-negative | 187 | 153 | 81.8% (75.7% - 86.7%) | 364 | 347 | 95.3% (92.6% - 97.1%) |
| YiDiXie™-SS in CA125-positive | 22 | 22 | 100% (85.1% - 100%) | 6 | 4 | 66.7% (30.0% - 94.1%) |
| YiDiXie™-HS in CA125-negative | 183 | 178 | 97.3% (93.8% - 98.8%) | 210 | 187 | 89.0% (84.1% - 92.6%) |
| YiDiXie™-D in CA125-positive | 22 | 22 | 100% (85.1% - 100%) | 6 | 6 | 100% (61.0% - 100%) |
| YiDiXie™-D in CA125-negative | 183 | 152 | 83.1% (77.0% - 87.8%) | 210 | 197 | 93.8% (89.7% - 96.3%) |
| YiDiXie™-SS in CA19-9-positive | 34 | 34 | 100% (89.8% - 100%) | 9 | 6 | 66.7% (35.4% - 87.9%) |
| YiDiXie™-HS in CA19-9-negative | 182 | 176 | 96.7% (93.0% - 98.5%) | 343 | 310 | 90.4% (86.8% - 93.1%) |
| YiDiXie™-D in CA19-9-positive | 34 | 34 | 100% (89.8% - 100%) | 9 | 9 | 100% (70.1% - 100%) |
| YiDiXie™-D in CA19-9-negative | 182 | 147 | 83.5% (77.4% - 88.2%) | 343 | 330 | 97.1% (94.7% - 98.4%) |

CI, confidence interval. <sup>ab</sup> Two-sided 95% Wilson confidence intervals were calculated.

### Diagnostic performance of YiDiXie™-HS in FOBT, CEA, CA125, CA19-9 negative patients

In order to solve the challenge of high false-negative rate of FOBT, CEA, CA125, CA19-9, YiDiXie™-HS was applied to FOBT, CEA, CA125, CA19-9 negative patients.

As shown in Table 5, YiDiXie™-HS substantially reduced the false-negative rate of FOBT, CEA, CA125, and CA19-9.

### Diagnostic performance of YiDiXie™-D in FOBT, CEA, CA125, CA19-9 positive patients

In order to further reduce the false-positive rates of FOBT, CEA, CA125, and CA19-9, YiDiXie™-D, which has relatively high sensitivity and very high specificity, was therefore applied.

As shown in Table 5, YiDiXie™-D substantially reduced the false positive rates of FOBT, CEA, CA125, and CA19-9.

### Diagnostic performance of YiDiXie™-D in FOBT, CEA, CA125, CA19-9 negative patients

In order to reduce the false-negative rates of FOBT, CEA, CA125, and CA19-9 while maintaining high specificity, YiDiXie™-D, which has relatively high sensitivity and very high specificity, was therefore applied.

As shown in Table 5, YiDiXie™-D significantly reduced the false-negative rates of FOBT, CEA, CA125, and CA19-9 while maintaining high specificity.

## DISCUSSION

### Clinical significance of YiDiXie™-SS in patients with positive FOBT and other indicators

The YiDiXie™ test includes three different test products: YiDiXie™-HS, YiDiXie™-SS, and YiDiXie™-D.^31^ Among them, YiDiXie™-HS combines sensitivity and specificity, with high sensitivity and high specificity.^31^ YiDiXie™-SS is highly sensitive to all malignant tumor types, but slightly less specific.^31^ YiDiXie™-D is highly specific to all malignant tumor types, but has low sensitivity^31^.

For patients with positive markers such as FOBT, the sensitivity and specificity of further diagnostic methods are very important. Weighing the contradiction between sensitivity and specificity is essentially weighing the contradiction between “the harm of missing diagnosis of malignant tumors” and “the harm of misdiagnosis of benign tumors”. In general, when markers such as FOBT are positive, gastroscopy is usually accepted, rather than radical surgery. Therefore, false positives of FOBT and other markers do not lead to serious consequences such as major surgical trauma, organ resection and loss of function. In this way, for patients with positive markers such as FOBT, the”harm of missed diagnosis of malignant tumor” is much higher than the “harm of misdiagnosis of benign tumor”. Therefore, YiDiXie ™ -SS with extremely high sensitivity but slightly low specificity was selected to reduce the false positive rate of markers such as FOBT.

As shown in Table 5, YiDiXie™-SS significantly reduced the false-positive rates of FOBT, CEA, CA125, and CA19-9 with essentially no increase in malignancy leakage. These results mean that YiDiXie™-SS significantly reduces the probability of erroneous gastroscope for benign gastric disease, with virtually no increase in missed diagnosis of malignant tumors.

In other words, YiDiXie™-SS significantly reduced mental distress, costly tests, and harm in patients with false-positive FOBT, CEA, CA125, and CA19-9 with virtually no increase in delayed treatment of malignant tumors. Therefore, YiDiXie™-SS well meets the clinical needs, has important clinical significance and wide application prospects.

### Clinical significance of YiDiXie™-HS in patients with negative FOBT and other indicators

For patients with negative markers such as FOBT, the sensitivity and specificity of further diagnostic methods are very important. Weighing the contradiction between sensitivity and specificity is essentially weighing the contradiction between “the harm of missing diagnosis of malignant tumors” and “the harm of misdiagnosis of benign diseases”. The higher false negative rate means that more malignant tumors are missed, which will lead to delayed treatment, the progression of malignant tumors, and even the development of advanced stage. Therefore, patients will have to bear the adverse consequences of poor prognosis, short survival, poor quality of life, and high treatment costs. A higher false positive rate means that more benign diseases are misdiagnosed, leading to unnecessarily expensive and invasive colonoscopy. As a result, patients have to bear the negative consequences of mental pain, expensive tests, and examination injuries. Therefore, YiDiXie ™ -HS with high sensitivity and specificity was selected to reduce the false-negative rate of FOBT and other markers.

As shown in Table 5, YiDiXie™-HS substantially reduced the false-negative rates of FOBT, CEA, CA125, and CA19-9. These results mean that YiDiXie™-HS significantly reduces the probability of false negative detection of malignant tumors by FOBT and other markers.

In other words, YiDiXie™-HS significantly reduces the adverse consequences of poor prognosis, high treatment costs, poor quality of life, and short survival for patients with false negative diagnosis of FOBT and other markers. Therefore, YiDiXie™-HS well meets the clinical needs and has important clinical significance and wide application prospects.

### Clinical Significance of YiDiXie™-D in Patients with Gastric Tumors

For patients with gastric tumors, YiDiXie™-D, which has relatively high sensitivity and very high specificity, can be used to further reduce the false-positive rates of FOBT, CEA, CA125, and CA19-9 or to significantly reduce their false-negative rates while maintaining high specificity.

As shown in Table 5, YiDiXie™-D substantially reduced the false-positive rates of FOBT, CEA, CA125, and CA19-9. YiDiXie™-D significantly reduced the false-negative rates of FOBT, CEA, CA125, and CA19-9 while maintaining a high degree of specificity.

The above results imply that YiDiXie ™ -D further reduces the risk of performing wrong gastroscopy for gastric tumors. Therefore, YiDiXie™ -D well meets the clinical needs and has important clinical significance and wide application prospects..

### YiDiXie™ tests promise to address 2 challenges in gastric cancer

First, YiDiXie ™ -SS can greatly relieve the unnecessary work pressure of digestive endoscopy physicians, and promote the timely diagnosis and timely treatment of malignant tumor cases that were originally delayed. When markers such as FOBT are positive, the patient usually receives a gastroscope. Whether these colonoscopies can be completed in a timely manner directly depends on the number of digestive endoscopists. In many parts of the world, reservations can take months or even more than a year. This inevitably delays the treatment of malignant tumor cases, so it is not uncommon to see malignant tumor progression or even distant metastasis in patients with positive markers such as FOBT waiting for colonoscopy.As shown in Table 5, YiDiXie™-SS significantly reduced the false-positive rates of FOBT, CEA, CA125, and CA19-9 with essentially no increase in malignancy leakage. Therefore, YiDiXie™-SS can greatly relieve the unnecessary work pressure of digestive endoscopists, and facilitate the timely diagnosis and treatment of gastric cancer or other diseases that have been delayed.

Second, YiDiXie™-HS significantly reduces the risk of missed gastric cancer. When FOBT and other markers are negative, the possibility of gastric cancer is usually temporarily ruled out. Due to the high false-negative rate of FOBT and other markers, a large number of gastric cancer patients have delayed treatment. As shown in Table 5, YiDiXie™ -HS substantially reduced the false-negative rates of FOBT, CEA, CA125, and CA19-9. These results mean that YiDiXie™-HS significantly reduces the probability of false negative detection of malignant tumors by FOBT and other markers. Therefore, YiDiXie™-HS significantly reduces the probability of false-negative missed malignant tumors by markers such as FOBT, and promotes timely diagnosis and treatment for gastric cancer patients who were previously delayed.

Again, YiDiXie ™ -D is expected to further address the challenges of “high false positive rate”and “high false negative rate “. As shown in Table 5, YiDiXie ™ -D significantly reduces the false-positive rates of FOBT, CEA, CA125, and CA19-9. YiDiXie ™ -D significantly reduces the false-negative rates of FOBT, CEA, CA125, and CA19-9 while maintaining a high degree of specificity. Thus, YiDiXie ™ -D further reduces the risk of incorrectly performing gastroscopy for gastric tumors.

Final, the YiDiXie™ test enables “just-in-time diagnosis” for gastric cancer patients. On the one hand, the YiDiXie ™ test requires only a tiny amount of blood, allowing patients to complete the diagnostic process without leaving their homes. The YiDiXie™ test requires only 20 μl of serum to complete, which is about the same amount as 1 drop of whole blood (1 drop of whole blood is about 50 μl, which produces 20-25μl of serum)^31^. Taking into account the sample quality assessment test before detection and 2-3 repetitions, 0.2 ml of whole blood was sufficient to complete the YiDiXie ™ test^31^. Ordinary subjects can use the finger blood collection needle to complete 0.2 ml finger blood collection at home, without the need for intravenous blood collection by medical personnel, and patients can complete the diagnosis process without leaving the house^31^.

The YiDiXie ™ test, on the other hand, has a nearly unlimited diagnostic capacity. The basic flow diagram of the YiDiXie™ test in Figure 1 shows that the YiDiXie ™ test not only does not require a doctor and medical equipment, but also does not require medical personnel to collect blood. As a result, the YiDiXie™ test is completely independent of the number of medical personnel and facilities, and its testing capacity is nearly unlimited. As a result, the YiDiXie ™ test enables “just-in-time diagnosis” for gastric cancer patients without the anxiety of waiting for an appointment.

**Figure 1.**
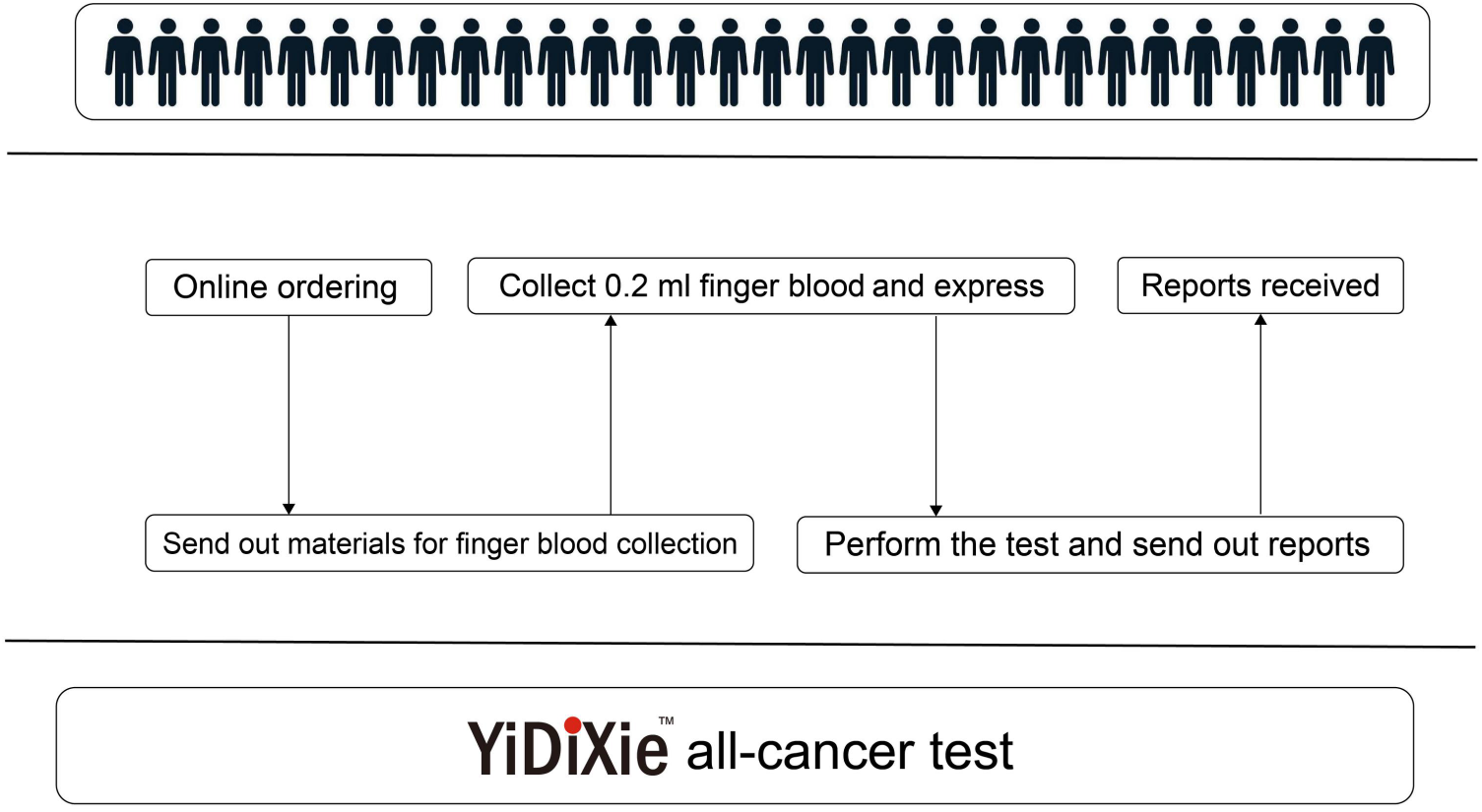
Basic flowchart of the YiDiXie™ test.

In short, the YiDiXie™ test has important diagnostic value in gastric cancer, which is expected to solve the two problems of “too high false-positive rate of FOBT and other markers” and “too high false-negative rate of FOBT and other markers” in gastric cancer.

### Limitations of the study

Firstly, the number of cases in this study is small, and future clinical studies with a larger sample size are needed for further evaluation.

Secondly, this study is a control study of inpatients with malignant tumors and benign tumors, and future cohort studies of natural lung tumors are needed for further evaluation.

Finally, this study is a single-center study, which may lead to a certain degree of bias in the results of this study. Future multi-center studies are needed for further evaluation.

## CONCLUSION

YiDiXie ™ -SS has very high sensitivity and relatively high specificity in gastric tumors.YiDiXie™ -HS has high sensitivity and high specificity in gastric tumors.YiDiXie ™ -D has relatively high sensitivity and very high specificity in gastric tumors. YiDiXie™-SS significantly reduces the false-positive rates of FOBT, CEA, CA125, and CA19-9 with essentially no increase in delayed treatment of gastric cancer.YiDiXie ™ -HS significantly reduces the false-negative rates of FOBT, CEA, CA125, and CA19-9.YiDiXie ™ -D can significantly reduce the false-positive rate of FOBT, CEA, CA125 and CA19-9, or significantly reduce the false-negative rate of FOBT, CEA, CA125 and CA19-9 while maintaining a high degree of specificity. YiDiXie ™ test has an important diagnostic value in gastric cancer, and is expected to solve the problems of “high false positive rate”and “high false negative rate”of FOBT, CEA, CA125 and CA19-9.

## Data Availability

All data produced in the present study are contained in the manuscript.

## FUNDING

This study was supported by Shenzhen High-level Hospital Construction Fund, Clinical Research Project of Peking University Shenzhen Hospital (LCYJ2020002, LCYJ2020015, LCYJ2020020, LCYJ2017001).\

## Notes

### Competing Interest Statement

The authors have declared no competing interest.

### Clinical Trial

ChiCTR2200066840

### Funding Statement

This work was supported by Shenzhen High-level Hospital Construction Fund, Clinical Research Project of Peking University Shenzhen Hospital (LCYJ2020002, LCYJ2020015, LCYJ2020020, LCYJ2017001).

### Author Declarations

Ethics committee of Peking University Shenzhen Hospital gave ethical approval for this work.

### Summary of Updates

The results were updated. Most tabels and results were reviesed.

## REFERENCES

1. Bray F, Laversanne M, Sung H, Ferlay J, Siegel RL, Soerjomataram I, Jemal A: Global cancer statistics 2022: GLOBOCAN estimates of incidence and mortality worldwide for 36 cancers in 185 countries. CA Cancer J Clin 2024, 74(3):229–263.

2. Sung H, Ferlay J, Siegel RL, Laversanne M, Soerjomataram I, Jemal A, Bray F: Global Cancer Statistics 2020: GLOBOCAN Estimates of Incidence and Mortality Worldwide for 36 Cancers in 185 Countries. CA Cancer J Clin 2021, 71(3):209–249.

3. Howson CP, Hiyama T, Wynder EL: The decline in gastric cancer: epidemiology of an unplanned triumph. Epidemiologic reviews 1986, 8:1–27.

4. Anderson WF, Rabkin CS, Turner N, Fraumeni JF, Jr., Rosenberg PS, Camargo MC: The Changing Face of Noncardia Gastric Cancer Incidence Among US Non-Hispanic Whites. Journal of the National Cancer Institute 2018, 110(6):608–615.

5. Arnold M, Park JY, Camargo MC, Lunet N, Forman D, Soerjomataram I: Is gastric cancer becoming a rare disease? A global assessment of predicted incidence trends to 2035. Gut 2020, 69(5):823–829.

6. Gu L, Khadaroo PA, Chen L, Li X, Zhu H, Zhong X, Pan J, Chen M: Comparison of Long-Term Outcomes of Endoscopic Submucosal Dissection and Surgery for Early Gastric Cancer: a Systematic Review and Meta-analysis. Journal of gastrointestinal surgery: official journal of the Society for Surgery of the Alimentary Tract 2019, 23(7):1493–1501.

7. Zhang XY, Zhang PY: Gastric cancer: somatic genetics as a guide to therapy. Journal of medical genetics 2017, 54(5):305–312.

8. He S, Xia C, Li H, Cao M, Yang F, Yan X, Zhang S, Teng Y, Li Q, Chen W: Cancer profiles in China and comparisons with the USA: a comprehensive analysis in the incidence, mortality, survival, staging, and attribution to risk factors. Science China Life sciences 2024, 67(1):122–131.

9. Mattiuzzi C, Lippi G: Current Cancer Epidemiology. Journal of epidemiology and global health 2019, 9(4):217–222.

10. Gao JP, Xu W, Liu WT, Yan M, Zhu ZG: Tumor heterogeneity of gastric cancer: From the perspective of tumor-initiating cell. World journal of gastroenterology 2018, 24(24):2567–2581.

11. Rockey DC, Koch J, Cello JP, Sanders LL, McQuaid K: Relative frequency of upper gastrointestinal and colonic lesions in patients with positive fecal occult-blood tests. The New England journal of medicine 1998, 339(3):153–159.

12. McCracken M, Olsen M, Chen MS, Jr., Jemal A, Thun M, Cokkinides V, Deapen D, Ward E: Cancer incidence, mortality, and associated risk factors among Asian Americans of Chinese, Filipino, Vietnamese, Korean, and Japanese ethnicities. CA Cancer J Clin 2007, 57(4):190–205.

13. Replogle ML, Glaser SL, Hiatt RA, Parsonnet J: Biologic sex as a risk factor for Helicobacter pylori infection in healthy young adults. American journal of epidemiology 1995, 142(8):856–863.

14. Everhart JE, Kruszon-Moran D, Perez-Perez GI, Tralka TS, McQuillan G: Seroprevalence and ethnic differences in Helicobacter pylori infection among adults in the United States. The Journal of infectious diseases 2000, 181(4):1359–1363.

15. Day LW, Cello JP, Somsouk M, Inadomi JM: Prevalence of gastric cancer versus colorectal cancer in Asians with a positive fecal occult blood test. Indian journal of gastroenterology: official journal of the Indian Society of Gastroenterology 2011, 30(5):209–216.

16. Ghosh I, Bhattacharjee D, Das AK, Chakrabarti G, Dasgupta A, Dey SK: Diagnostic Role of Tumour Markers CEA, CA15-3, CA19-9 and CA125 in Lung Cancer. Indian journal of clinical biochemistry: IJCB 2013, 28(1):24–29.

17. Huang G, Chen R, Lu N, Chen Q, Lv W, Li B: Combined Evaluation of Preoperative Serum CEA and CA125 as an Independent Prognostic Biomarker in Patients with Early-Stage Cervical Adenocarcinoma. OncoTargets and therapy 2020, 13:5155–5164.

18. Zhang J, Wei Q, Dong D, Ren L: The role of TPS, CA125, CA15-3 and CEA in prediction of distant metastasis of breast cancer. Clinica chimica acta; international journal of clinical chemistry 2021, 523:19–25.

19. Dolscheid-Pommerich RC, Manekeller S, Walgenbach-Brünagel G, Kalff JC, Hartmann G, Wagner BS, Holdenrieder S: Clinical Performance of CEA, CA19-9, CA15-3, CA125 and AFP in Gastrointestinal Cancer Using LOCI™-based Assays. Anticancer research 2017, 37(1):353–359.

20. Ruibal Morell A: CEA serum levels in non-neoplastic disease. The International journal of biological markers 1992, 7(3):160–166.

21. Kornek GV, Depisch D, Rosen HR, Temsch EM, Scheithauer W: Comparative analysis of CA72-4, CA195 and carcinoembryonic antigen in patients with gastrointestinal malignancies. Journal of cancer research and clinical oncology 1992, 118(4):318–320.

22. Lordick F, Carneiro F, Cascinu S, Fleitas T, Haustermans K, Piessen G, Vogel A, Smyth EC: Gastric cancer: ESMO Clinical Practice Guideline for diagnosis, treatment and follow-up. Annals of oncology: official journal of the European Society for Medical Oncology 2022, 33(10):1005–1020.

23. Association: Japanese Gastric Cancer Treatment Guidelines 2021 (6th edition). Gastric cancer: official journal of the International Gastric Cancer Association and the Japanese Gastric Cancer Association 2023, 26(1):1–25.

24. Ajani JA, D’Amico TA, Bentrem DJ, Chao J, Cooke D, Corvera C, Das P, Enzinger PC, Enzler T, Fanta P et al: Gastric Cancer, Version 2.2022, NCCN Clinical Practice Guidelines in Oncology. Journal of the National Comprehensive Cancer Network: JNCCN 2022, 20(2):167–192.

25. Bond JH: FOBT is not an effective way to screen for gastric cancer. Digestive and liver disease: official journal of the Italian Society of Gastroenterology and the Italian Association for the Study of the Liver 2007, 39(4):327–328.

26. Park SH, Ku KB, Chung HY, Yu W: Prognostic significance of serum and tissue carcinoembryonic antigen in patients with gastric adenocarcinomas. Cancer research and treatment 2008, 40(1):16–21.

27. Wang W, Chen XL, Zhao SY, Xu YH, Zhang WH, Liu K, Chen XZ, Yang K, Zhang B, Chen ZX et al: Prognostic significance of preoperative serum CA125, CA19-9 and CEA in gastric carcinoma. Oncotarget 2016, 7(23):35423–35436.

28. Liang Y, Wang W, Fang C, Raj SS, Hu WM, Li QW, Zhou ZW: Clinical significance and diagnostic value of serum CEA, CA19-9 and CA72-4 in patients with gastric cancer. Oncotarget 2016, 7(31):49565–49573.

29. Feng F, Tian Y, Xu G, Liu Z, Liu S, Zheng G, Guo M, Lian X, Fan D, Zhang H: Diagnostic and prognostic value of CEA, CA19-9, AFP and CA125 for early gastric cancer. BMC cancer 2017, 17(1):737.

30. He CZ, Zhang KH, Li Q, Liu XH, Hong Y, Lv NH: Combined use of AFP, CEA, CA125 and CAl9-9 improves the sensitivity for the diagnosis of gastric cancer. BMC gastroenterology 2013, 13:87.

31. Chen Sun, Chong Lu, Yongjian Zhang, Ling Wang, Zhenjian Ge, Zhenyu Wen, Wenkang Chen, Yingqi Li, Yutong Wu, Shengjie Lin et al: Evaluation of the Multi-Cancer Early Detection (MCED) value of YiDiXie ™-HS and YiDiXie™-SS. medRxiv 2024:doi: 10.1101/2024.1103.1111.24303683.

32. Edge SB, Compton CC: The American Joint Committee on Cancer: the 7th edition of the AJCC cancer staging manual and the future of TNM. Ann Surg Oncol 2010, 17(6):1471–1474.

33. Amin MB, Greene FL, Edge SB, Compton CC, Gershenwald JE, Brookland RK, Meyer L, Gress DM, Byrd DR, Winchester DP: The Eighth Edition AJCC Cancer Staging Manual: Continuing to build a bridge from a population-based to a more “personalized” approach to cancer staging. CA Cancer J Clin 2017, 67(2):93–99.

